# DNA methylation partially mediates the relationship between childhood adversity and depressive symptoms in adolescence

**DOI:** 10.1101/2021.06.28.21259426

**Authors:** Brooke J. Smith, Alexandre A. Lussier, Janine Cerutti, Andrew J. Simpkin, Andrew D.A.C. Smith, Matthew J. Suderman, Esther Walton, Daniel J. Schaid, Erin C. Dunn

## Abstract

**Background:** Exposure to adversity during childhood is estimated to at least double the risk of depression later in life. Some evidence suggests childhood adversity may have a greater impact on depression risk, if experienced during specific windows of development called sensitive periods. During these sensitive periods, there is evidence that adversity may leave behind biological memories, including changes in DNA methylation (DNAm). Here we ask if those changes play a role in the link between adversity and later adolescent depressive symptoms.

**Methods:** We applied a method for high-dimensional mediation analysis using data from a subsample (n=627-675) of the Avon Longitudinal Study of Parents and Children. We first assessed the possibility of time-dependent relationships between seven types of childhood adversity (caregiver abuse, physical/sexual abuse, maternal psychopathology, one-adult household, family instability, financial stress, neighborhood disadvantage), measured on at least four occasions between ages 0-7 years, and adolescent depression at mean age 10.6. Specifically, we considered three types of life course hypotheses (sensitive periods, accumulation, and recency), and then evaluated which of these hypotheses had the strongest association in each adversity-adolescent depression relationship using the structured life course modeling approach (SLCMA; pronounced “slick-mah”). To conduct the mediation analyses, we used a combination of pruning and sure independence screening (a dimension reduction method) to reduce the number of methylated CpG sites under consideration to a viable subset for our sample size. We then applied a sparse group lasso penalized model to identify the top mediating loci from that subset using the combined strength of the coefficient measuring the relationship between the childhood adversity and a CpG site (*α*) and of the coefficient measuring the relationship between the CpG site and depressive symptoms (*β*) as a metric. Using a Monte Carlo method for assessing mediation (MCMAM), we assigned a significance level and confidence interval to each identified mediator.

**Results:** Across all seven adversities, we identified a total of 70 CpG sites that showed evidence of mediating the relationship between adversity and adolescent depression symptoms. Of these 70 mediators, 37 were significant at the p < 0.05 level when applying the MCMAM, a method tailored to estimating the significance of SEM-derived mediation effects. These sites exhibited four different mediating patterns, differentiated by the direction of *α* and *β*. These patterns had signals that were: (1) both positive (19 loci), (2) both negative (18 loci), (3) positive *α* and negative *β* (23 loci) or (4) negative *α* and positive *β* (10 loci).

**Conclusion:** Our results suggest that DNAm partially mediates the relationship between different types of childhood adversity and depressive symptoms in adolescence. These findings provide insight into the biological mechanisms that link childhood adversity to depression, which will ultimately help develop treatments to prevent depression in more vulnerable populations.

## Introduction

Depression is one of our greatest health problems, affecting over 264 million people globally (1). Within the next ten years, depression is projected to inflict a greater burden than any other disease in the world (2). Yet despite its prevalence and impact, relatively little is known about the etiology of depression and therefore how to prevent it.

From decades of research, exposure to childhood adversity (e.g., abuse, violence) has emerged as one of the strongest and most consistent contributors to risk for depression and other psychiatric disorders (3). Indeed, a recent meta-analysis found that nearly half of all people with depression report having experienced childhood maltreatment (4). If the associations between childhood adversities and later depression are causal, childhood adversity could explain up to one-third of all psychiatric disorders (5). However, the biological pathways through which childhood adversity contributes to disease risk are poorly understood.

A reasonable hypothesis is that these negative early-life exposures take hold through epigenetic changes. DNA methylation (DNAm) is an epigenetic mechanism in which methyl groups are added to DNA, potentially altering gene expression without any changes to the actual DNA sequence (6). The majority of DNA methylation occurs on cytosines that precede a guanine nucleotide, called CpG sites. The consensus from systematic reviews is that different types of childhood adversity can impact DNAm levels (7, 8). Recent work suggests some types of childhood adversity might have their greatest effects on DNAm levels during early childhood, and specifically during a sensitive period between ages 3 to 5 years (9, 10). Furthermore, DNAm patterns have been linked, primarily in samples of adults, to future depression and depressive symptoms (11-14). For instance, baseline DNAm profiles in patients diagnosed with major depressive disorder (MDD) predicted their disease course six years later, suggesting DNAm may serve as a valuable biomarker for response to treatment or symptom changes (15). Even more intriguing, recent experimental studies in humans have shown that psychiatric interventions can *reverse* DNAm signatures previously associated with psychopathology (16-18).

However, a key question remains unanswered: is DNAm the bridge between these social environmental experiences and risk for depression? Studies using mediation analysis are uniquely poised to answer this question. Mediation analysis determines whether an association between an exposure and outcome persists, at least in part, through an intermediate variable or “mediator”. A mediator is both a consequence of the exposure *and* an influence on the outcome. Using mediation analysis, we will investigate the hypothesis that DNAm mediates the effect of childhood adversity on later depression risk. If DNAm is a causal intermediary of the adversity-psychopathology relationship, then it may be possible to identify individuals at higher risk for psychopathology based on their epigenome and ultimately, reduce the overall burden of mental illness.

To our knowledge, no studies to date have investigated prospectively whether the relationship between childhood adversity and later depression symptoms is mediated by genome-wide DNAm changes (19). However, several studies have examined DNAm as a mediator between similar environmental exposures and biological health outcomes, such as between environmental risks like maternal psychopathology and callous-unemotional traits in youth (20), between childhood trauma and stress cortisol reactivity (21), and between prenatal smoking and substance-use risk (22).

The goal of this paper is to investigate whether DNAm mediates the relationship between common types of childhood adversity and depressive symptoms in early adolescence. To conduct these analyses, we used data from the Avon Longitudinal Study of Parents and Children (ALSPAC), a longitudinal birth cohort that uniquely had repeated measures of childhood adversity exposure in addition to DNAm data and measures of depressive symptoms during early adolescence.

## Methods

### Sample and Procedures

Our data source, ALSPAC, is a 30-year-long ongoing prospective birth cohort study that recruited 14,541 pregnant women residing in Avon, UK, with expected delivery dates between 1 April 1991 and 31 December 1992 (23, 24). Further details of the study and available data are provided on the study website through a fully searchable data dictionary (http://www.bris.ac.uk/alspac/researchers/data-access/data-dictionary/). All data are available by request from the ALSPAC Executive Committee for researchers who meet the criteria for access to confidential data (http://www.bristol.ac.uk/alspac/researchers/access/). Ethical approval for the study was obtained from the ALSPAC Law and Ethics Committee and the Local Research Ethics Committees. Consent for biological samples was collected in accordance with the Human Tissue Act (2004). Informed consent for the use of data collected via questionnaires and clinics was obtained from participants following the recommendations of the ALSPAC Ethics and Law Committee. Secondary analyses of these data were approved with oversight by the Mass General Brigham Institutional Review Boards (IRB) (Protocol 2017P001110).

We analyzed data specifically from the Accessible Resource for Integrated Epigenomic Studies (ARIES), a subset of 1,018 mother-child pairs who were randomly selected from ALSPAC based on having DNA samples across at least five waves (two for the mother, three for the child). For our analyses, we looked specifically at singleton birth children who had complete data on exposure, outcome, and all covariates of interest.

### Measures

#### Exposure to Adversity

We examined seven types of commonly-occurring childhood adversities: 1) caregiver emotional or physical cruelty, 2) sexual or physical abuse (by anyone), 3) maternal psychopathology, 4) financial stress, 5) family instability, 6) one-adult households, and 7) neighborhood disadvantage. These adversities were selected because they have previously been associated in numerous studies with epigenetic marks (25-28), and elevated risk for both depression and internalizing symptoms in youth (29, 30). Each adversity was reported by mothers using mailed questionnaires on at least four occasions from birth to age seven years.

Details on the specific times and measures used to derive each adversity can be found in

### Supplemental Materials

#### DNA methylation

As described elsewhere (31), DNAm was measured at 485,000 CpG sites across the genome using the Illumina Infinium Human Methylation 450K BeadChip microarray (Illumina, San Diego, CA). The DNA used for this assay was collected from whole blood and peripheral blood leukocytes when children were seven years old. Extractions were completed within three weeks after DNA collection (32). The University of Bristol performed the DNAm wet laboratory procedures, preprocessing analyses, and quality control (31, 33). DNAm values were expressed using beta values, which represent the proportion of methylated cells at each observed CpG site. DNAm data were processed using a pipeline based on the *meffil* R package developed by Min and colleagues (33). The pipeline included background correction and functional normalization to diminish variation due to technical artifacts. Furthermore, samples with more than 10% missing CpG site measurements (detection p-value > 0.01 or bead count < 3) were removed. Cross-hybridizing probes and polymorphic probes were removed leaving 450,745 probes.

To reduce the impact of potential outliers, we winsorized DNAm values for each CpG site by setting the bottom 5% and top 95% to the values for the 5^th^ and 95^th^ quantiles, respectively (34). We controlled for cell-type heterogeneity by estimating cell counts from DNAm measurements using the Houseman method (35) and then regressed the cell count estimates from our DNAm measurements as advised by Edgar and colleagues (36). Finally, we converted these adjusted DNAm measurements, represented as beta values (values between 0 and 1 indicating the ratio of methylated to unmethylated cells), to *M* values (the log_2_-transformation) because, unlike beta values, *M* values do not suffer from severe heteroskedasticity (unequal variance of the residuals) and therefore provide a more accurate detection and true positive rate than beta values (37). However, beta values were used in the plotting of all graphs as they have a more intuitive biological interpretation (i.e., percent of methylated cells).

#### Depressive Symptoms

Depressive symptoms at a mean age of 10.6 years were measured using scores derived from the child-completed Short Mood and Feelings Questionnaire (SMFQ), a frequently-used tool in population-based studies, which captures depressive symptoms in children and adolescents (38, 39). Children completed the SMFQ during a half-day session they attended at the ALSPAC clinic site. The 13-item measure uses a 3-point Likert scale with values 0=not true, 1=sometimes true, and 2=true to capture the child’s mental state over the previous two weeks.

Items include: “I felt miserable or unhappy”, “I felt lonely”, and “I cried a lot” (40). Further details about the SMFQ and its reliability can be found in **Supplemental Materials**. We focused on total SMFQ scores, rather than a binary indicator for probable depression, because continuous scores enable detection of more subtle variability in the data, which leads to greater precision and improved statistical power.

Being a longitudinal study, ALSPAC had repeated measures of depressive symptoms across time. We focused on the age 10 timepoint for several reasons. First, it was the next measurement of childhood depressive symptoms that occurred after the DNAm assessment at age 7 years. Thus, we could maintain temporality in the association between exposure to adversity prior to age 7, DNAm at age 7, and subsequent depressive symptoms, reducing concerns about reverse causation. Furthermore, in large population-based samples like ALSPAC, reports of child depressive symptoms are rarer before age 10. Indeed, age 10 is the first occasion in ALSPAC when children self-reported their own levels of depressive symptoms. This measure was advantageous because self-reports of depressive symptoms are often more reliable than parental reports, which typically underreport depressive symptoms (41). Therefore, child self-reports provide a more adequate measure of actual depressive symptoms and avoid biases resulting from parental reports (42). Age 10 was also advantageous as it allowed us to observe more immediate effects of DNAm changes on depressive symptoms and do so before the onset of puberty. Interpretation of the effects of DNAm on depressive symptoms become more complicated after puberty, when high-risk behaviors like smoking and drinking develop. Finally, at this age, self-reports of depression begin diverging from parental reports

#### Covariates

We controlled for several sociodemographic characteristics that have been linked to differences in DNAm among our study population and others and may potentially confound the associations in our analyses (43, 44). These sociodemographic covariates were child sex, child birthweight, child race and/or ethnicity, maternal age at gestation, maternal education level at gestation, number of previous pregnancies, and sustained maternal smoking during pregnancy (see **Supplemental Materials** for additional details).

### Data Analysis

We performed univariate and bivariate analyses of all covariates and our seven different exposures to adversity, assessing differences between groups using chi-squared and t-tests.

Following these initial analyses, we analyzed data in six main steps, as shown in **Figure 1A**.

**Figure 1.**
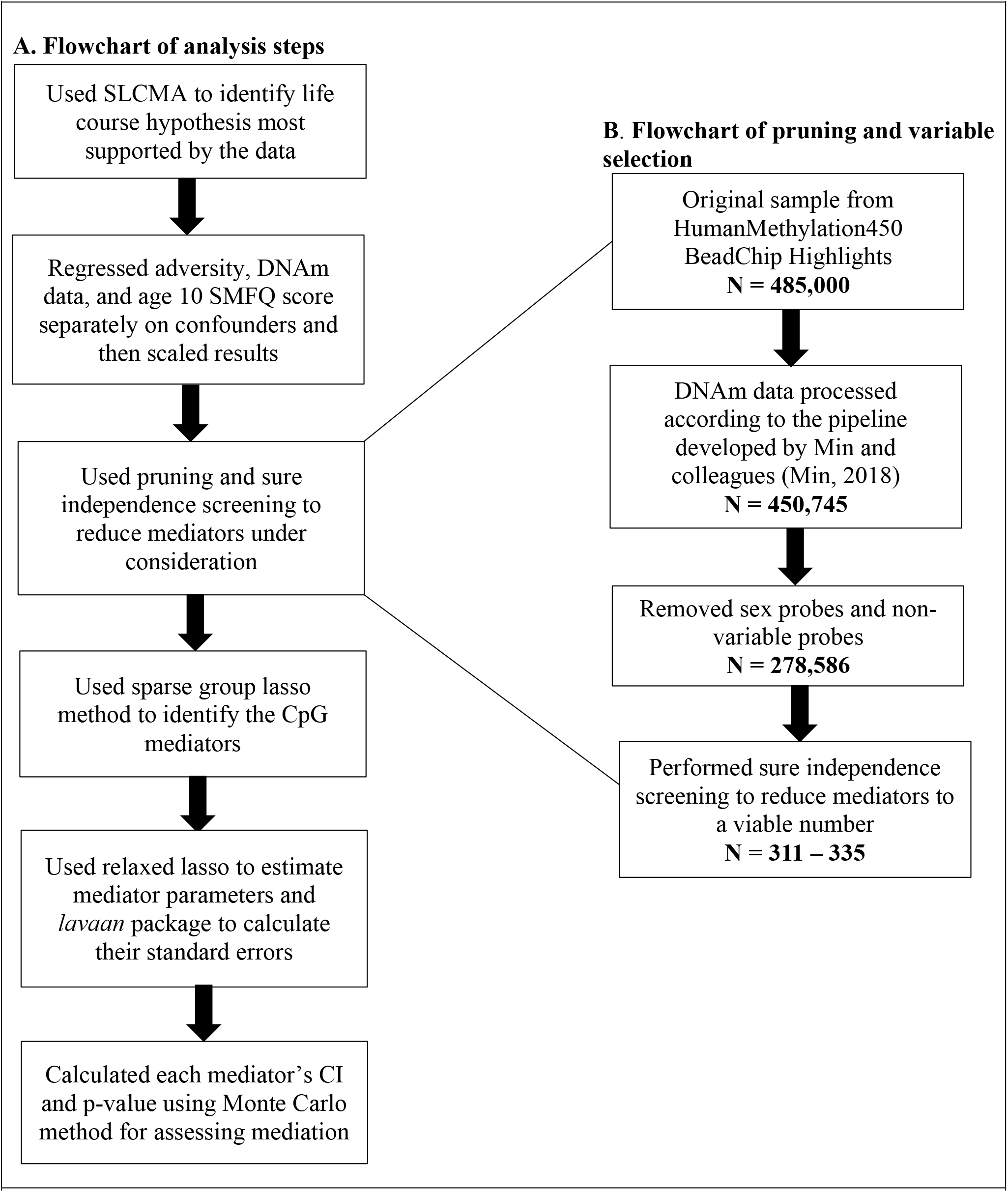
Flowchart of analysis steps for each adversity-outcome relationship. The analysis was performed seven times, once for each childhood adversity. **1A**. depicts the steps of the mediation analysis performed seven times, once for each of the seven adversities we studied. **1B**. highlights the steps to narrow down the mediators under consideration to a viable number for each of the seven adversities.

#### Selecting the best explanatory life course exposure

First, we used the structured life course modeling approach (SLCMA; pronounced “slick-mah”) to identify the life course hypothesis for each adversity that explained the most variation in early adolescent depressive symptoms. In brief, the SLCMA is a two-stage modeling approach that enables researchers to compare multiple life course hypotheses and identify the most parsimonious explanation for outcome variation (45-47). In the first stage, a variable selection tool called least angle regression (LARS) selects the life course model with the highest explanatory power for inclusion in the statistical model. In the second stage, post-selection inference is applied to calculate the total effect of adversity on depressive symptoms for the selected life course model. This modeling approach provides a distinct advantage over simpler modeling techniques like linear regression because it allows for consideration of multiple life course hypotheses *a priori* and bases selection of the life course hypothesis on its explanatory power rather than on hypotheses developed after observation. The SLCMA also allows for more nuanced life course modeling that considers timing and frequency of exposure unlike a simplistic ever-exposed hypothesis.

We considered five life-course hypotheses in the SLCMA: 1) *very early childhood sensitive period*, which tested whether adversity had an outsized impact during ages 0-2 years, 2) *early childhood sensitive period*, which tested during ages 3-5 years, 3) *middle childhood sensitive period*, which tested during ages 6-7 years, 4) *accumulation*, which measured whether the effects of repeated adversity accumulate over the life course (with individuals at higher accumulation experiencing larger effects), and 5) *recency*, in which effects of adversity were treated cumulatively but weighted by the age at which the adversity occurred. The single best-fitting life course model selected by the SLCMA for each adversity was then carried forward to subsequent mediation analyses. See **Supplemental Materials** for details on how the exposure variables were parameterized in the SLCMA.

#### Controlling for confounding and scaling

Second, to control for confounding and help sustain our power, we regressed the SLCMA-selected exposure hypothesis, DNAm data, and age 10 SMFQ score separately on the previously mentioned covariates. We then scaled these model inputs so that our results could be interpreted in terms of correlation between childhood adversity and depressive symptoms.

#### High dimensional mediation analysis

Using statistical mediation analyses, we determined the extent to which DNAm, as measured through individual CpG sites, mediated the relationship between each type of childhood adversity and adolescent depressive symptoms (**Supplemental Figure 2**).

**Figure 2.**
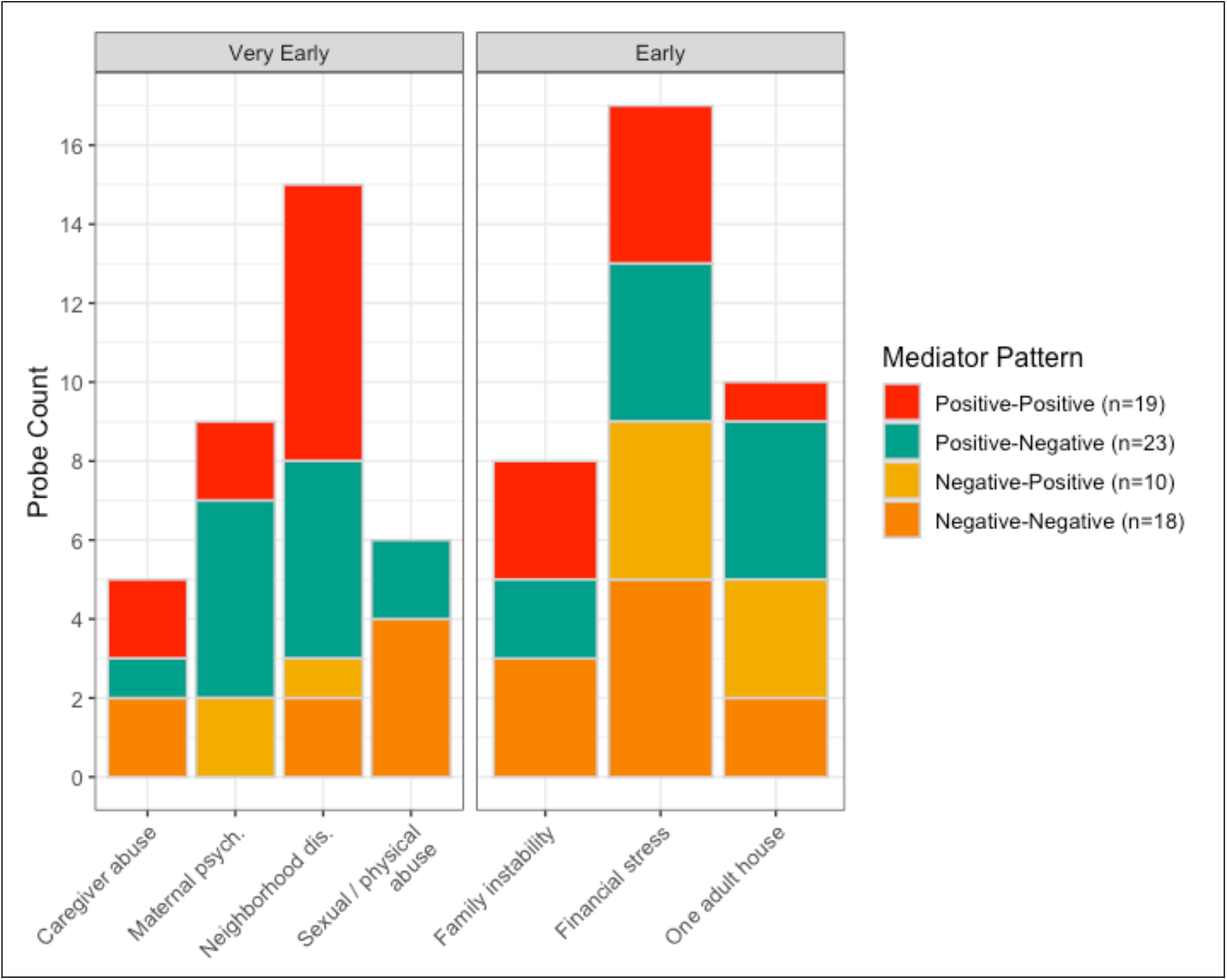
Mediator pattern by sensitive period model and type of adversity. Panels divide adversities by the theoretical sensitive period model under which they were selected, either “very early” or “early”. Colors indicate mediator pattern. Of the 70 mediators identified, 35 were associated with a very early sensitive period and 35 with an early sensitive period.

As described elsewhere (48, 49), testing for mediation using DNAm data is particularly challenging due to the high-dimensional nature of possible mediators. Our analytic samples consisted of all children with complete cases (n=627 to 675), meaning that the number of potential mediators (450,745 CpGs) was over 650 times greater than the number of observations. Therefore, as our third step before conducting the mediation analyses, we used pruning and sure independence screening (SIS), a method applied widely for dimension reduction in big data settings (50), to reduce the number of potential DNAm mediators to a viable number, *q* = ⌊(*n* - 3 - 1)/2⌋, (**Figure 1B**). Using SIS, we chose the top *q* CpG sites based on which had the highest correlation with both exposure and outcome. Further details as to how CpG sites were pruned and reduced using SIS can be found in **Supplemental Materials**.

#### The sparse group lasso penalized model

Fourth, we used a structural equation modeling approach developed by Schaid and Sinnwell called the sparse group lasso penalized model (hereto, referred to as the “Schaid-Sinnwell model”) to conduct the seven mediation analyses (51). The Schaid-Sinnwell model aims to optimize the regression model’s ability to measure mediation with the minimal number of mediators. Structural equation models (SEMs) are optimal for situations with multiple mediators, because they can simultaneously model the relationship among variables, account for the correlation between mediators, and estimate the direct effect (the effect of the childhood adversity on depression, independent of the influence of CpG site DNAm as an intermediate variable) and indirect effect (the portion of the effect mediated by CpG site DNAm between childhood adversity and depression) all within a single model (52-54).

The Schaid-Sinnwell model is particularly well-suited for mediation analysis as it groups the effect estimates for each mediator (i.e., association of exposure with mediator and association of mediator with outcome), rather than considering them separately, and it encourages sparseness (meaning parsimony) of the parameters by using a penalty (shrinkage) parameter, λ. A full description of the benefits of using the Schaid-Sinnwell model, as well as further details on its specific methodology can be found elsewhere (51). Additional details on its use in our analyses can be found in **Supplemental Materials**.

#### Estimating effect estimates and standard errors

Fifth, as penalized models tend to excessively shrink parameter estimates (51, 55), we used the relaxed lasso approach to calculate our effect sizes. This approach refits the selected model parameters without constraining them. We used the R package *lavaan* to approximate standard errors using the *sem* function (56).

#### Monte Carlo for p-value and confidence interval estimation

Finally, we assessed the statistical significance of our results by estimating a p-value and confidence interval of the indirect effect for each CpG mediator using the Monte Carlo method for assessing mediation (MCMAM) (57). Of note, we can estimate the indirect effect of a single mediator among multiple mediators because we used a SEM for our mediation analysis, which ensured that all mediators were regressed on one another.

## Results

### Sample demographics

Demographic information for the largest analytic sample – caregiver physical or emotional abuse – is shown in **Supplemental Table 2**. The analytic sample did not differ significantly from the ARIES sample. The ARIES sample, however, differed from the full ALSPAC cohort on all demographic variables except sex and SMFQ score. Most of these differences were slight in magnitude, except for maternal smoking (ARIES: 10.8% vs. ALSPAC: 21.2%) and maternal education, in which the ARIES sample contained mothers with higher education levels.

In our analytic sample, sexual or physical abuse (by anyone) had the lowest prevalence at 3.3% and maternal psychopathology had the highest at 13.5% (**Supplemental Figure 1**).

Depressive symptom scores averaged 3.70 (SD: 3.27) out of a possible score of 20 and did not statistically differ between the three samples.

### SLCMA results

Using the SLCMA, we found that exposure to adversity during sensitive periods in very early childhood (birth through age 2 years) and early childhood (ages 3-5 years) explained the most variation in depressive symptoms at age 10 years (**Table 1**). These sensitive periods of adversity were used as exposure measurements in the subsequent mediation analyses.

**Table 1.** Results of Structured Life Course Modeling Approach (SLCMA).

| **Adversity** | **Timepoint**<br>**Selected** | **Model Selected** | **Total**<br>**Effect** | **SE** | **p-value** | **CI** |
| --- | --- | --- | --- | --- | --- | --- |
| Caregiver physical or emotional abuse | 8 months | Very early childhood (0-3) | -0.81 | 0.64 | 0.90 | (-1.27, 36.45) |
| Sexual or physical abuse (by anyone) | 18 months | Early childhood (3-5) | 1.81 | 0.70 | 0.93 | (-Inf, 1.52) |
| **Maternal psychopathology** | **33 months** | **Very early childhood (0-3)** | **1.12** | **0.38** | **0.02** | **(0.07, 1.86)** |
| One adult in the household | 47 months | Early childhood (3-5) | 0.91 | 0.57 | 0.39 | (-3.80, 1.95) |
| Family instability | 57 months | Early childhood (3-5) | 1.21 | 0.68 | 0.80 | (-23.33, 1.93) |
| Financial Stress | 61 months | Early childhood (3-5) | 1.24 | 0.53 | 0.92 | (-Inf, 1.23) |
| Neighborhood disadvantage | 21 months | Very early childhood (0-3) | 1.04 | 0.48 | 0.07 | (-0.40, 1.99) |

Results of the SLCMA to identify the theoretical life course model with the best explanatory power between each adversity and the outcome of depressive symptoms in adolescence as measured by SMFQ score at age 10. For all adversities, a sensitive period was selected. We show the timepoint selected, its corresponding sensitive period model, the effect estimate for this sensitive period exposure on SMFQ score along with the standard error, p-value, and confidence interval. -Inf meaning negative infinity, can occur when samples are on the smaller end. CI = 95% confidence intervals; SE = standard error.

**Table 2.** Mediators selected by type of childhood adversity.

| **Adversity** | **Sample size** | **No. Mediators** | **Indirect effect** | **Direct Effect** | **Total Effect** |
| --- | --- | --- | --- | --- | --- |
| Caregiver physical or emotional abuse | 675 | 5 | 0.013 | -0.062 | -0.049 |
| Sexual or physical abuse | 663 | 6 | -0.073 | 0.173 | 0.100 |
| Maternal psychopathology | 653 | 9 | -0.026 | 0.143 | 0.117 |
| One adult in the household | 667 | 10 | -0.044 | 0.106 | 0.062 |
| Family instability | 656 | 8 | 0.007 | 0.061 | 0.068 |
| Financial stress | 627 | 17 | 0.028 | 0.065 | 0.093 |
| Neighborhood disadvantage | 660 | 15 | -0.011 | 0.093 | 0.082 |

Mediators selected for each adversity with measures of the total indirect, direct, and total effects summed by adversity.

### Mediation results summary

#### Schaid-Sinnwell model results

We identified 70 CpG sites across all seven adversities that showed evidence of being mediators; these sites were identified from the seven subsets ranging from 311 to 335 sites identified in the fourth analytic step. The penalized model selected CpG sites based on the best-fitting model as evaluated by the Bayesian Information Criterion (BIC), a method that avoids model over-fitting. Of the 70 sites, three sites (cg10953317, cg24059871, cg22239534) were selected in two different adversities, meaning 67 unique CpG sites were identified overall.

Thirty-seven sites were significant at the p<0.05 level.

**Table 2** provides a breakdown of the number of mediation sites found, as well as the estimated total indirect effect (the portion of the effect between childhood adversity and depressive symptoms mediated by all the selected CpG mediators), direct effect (the effect of the childhood adversity on depressive symptoms, independent of the influence of the selected CpG mediators), and total effect per adversity (sum of the direct and total indirect effects). The number of mediators per adversity ranged from 5 to 17. Four adversities had a negative total indirect effect, indicating that childhood adversity led to changes in DNAm that reduced depressive symptoms, and a positive direct effect, indicating that childhood adversity increased depressive symptoms independently of DNAm. In all but one adversity, the total effect – the sum of direct and indirect effects – was positive, indicating that childhood adversity still increased depressive symptoms despite the mediating effect of DNAm. Because childhood adversity, DNAm, and depressive symptom data were standardized prior to the mediation analyses, these effect estimates can be interpreted as partitioning the *amount of correlation explained* between exposure and outcome. Across all adversity mediation models, the percent of the Pearson’s partial correlation explained ranged from r=0.05-0.12 (mean=0.08).

#### Mediation patterns for individual CpG sites

Here, we use *α* to denote the effect of adversity on DNAm levels and *β* to denote the effect of DNAm on depressive symptoms (**Supplemental Figure 2**). The product *αβ* is the indirect effect of adversity on depressive symptoms mediated by DNAm.

The 70 CpG sites identified exhibited a mix of four different patterns based on the signs of the *α* and *β* coefficients described above (**Figure 2**): *positive-positive* (a positive *α* and *β*), *positive-negative* (a positive *α* and a negative *β*), *negative-positive* (a negative *α* and a positive *β*), and *negative-negative* (a negative *α* and *β*). There was no statistical difference in distribution of patterns across adversities (p=0.2). There were 35 sites selected for the very early childhood sensitive period and 35 sites selected for the early childhood sensitive period.

The *positive-positive* pattern was found for 19 CpG sites overall (**Figure 3A**). Here, children who were exposed to childhood adversity during the sensitive period had higher DNAm levels at the given CpG, which in turn, were correlated with increased depressive symptoms. CpGs exhibiting this pattern explained r=0.01-0.05 of the correlation between childhood adversity and depressive symptoms.

**Figure 3.**
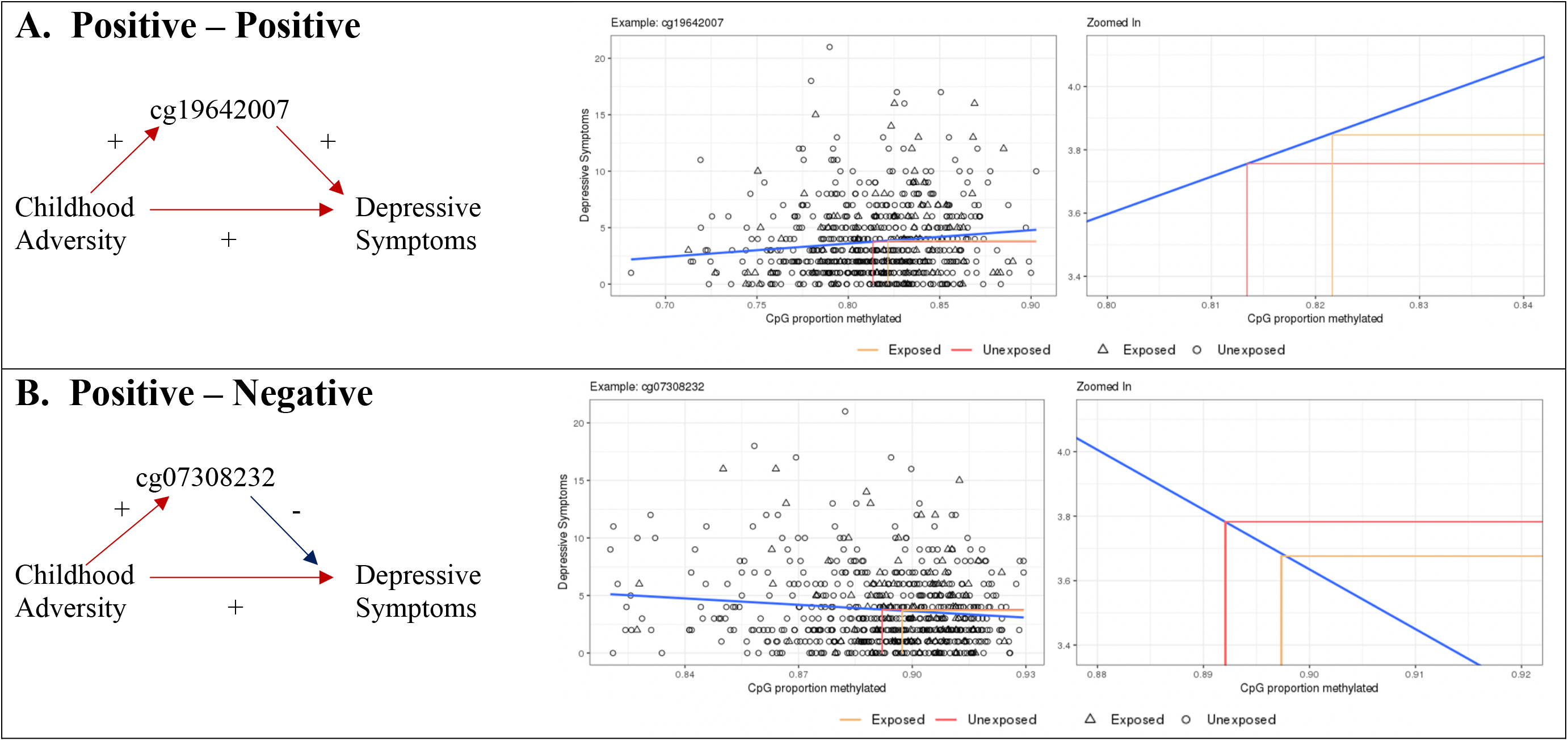
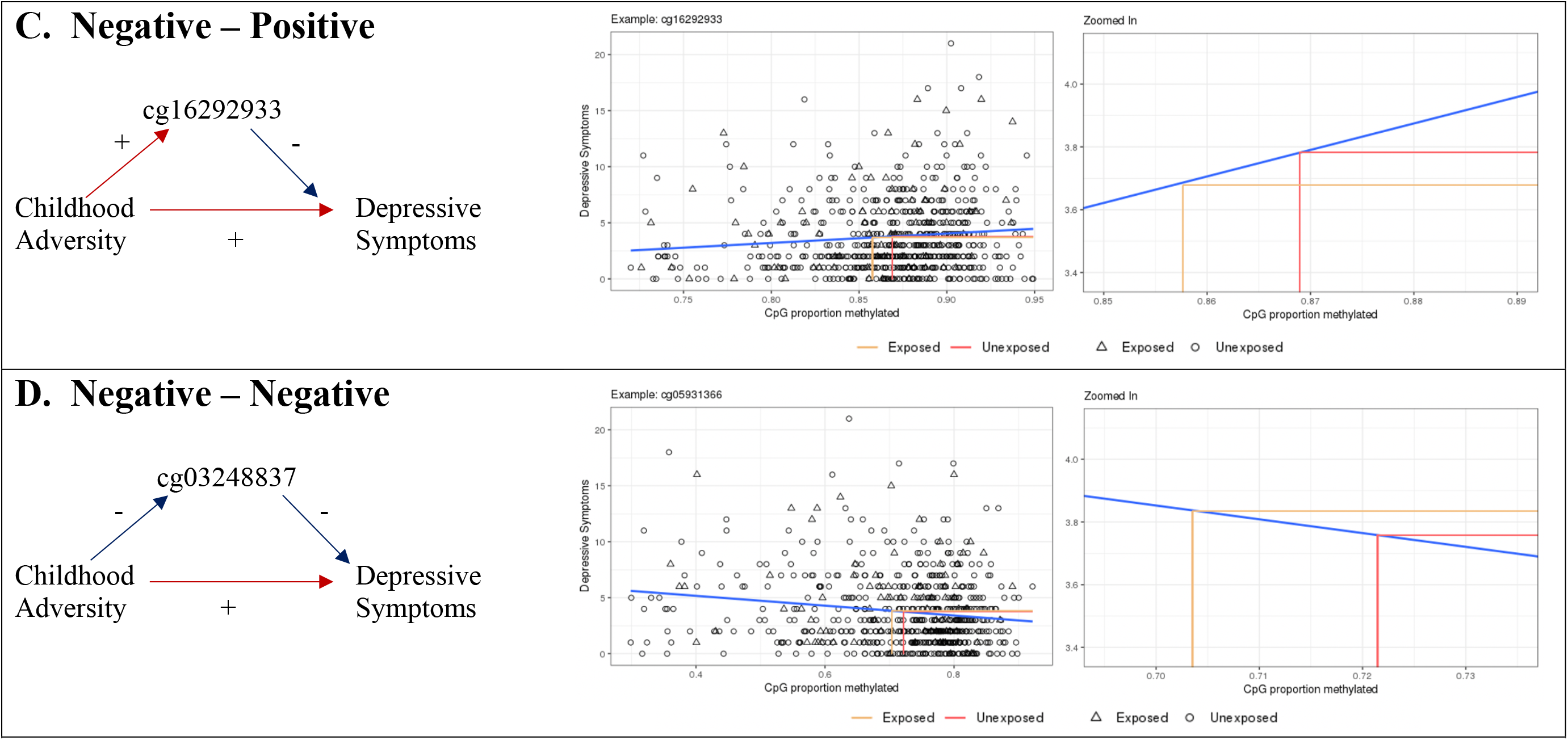
Mediation patterns for individual CpG sites. Panels A-D depict a mediating pattern. The directed acyclic graph (DAG) on the left illustrates how the adversity relates to the CpG mediator and how the CpG mediator relates to depressive symptoms. Each panel features a scatterplot containing all data points in the sample, accompanied by a best fit line. Pink and orange lines represent mean values for unexposed vs. exposed, respectively. In each zoomed-in plot, where the pink and orange lines meet the x-axis reflects mean DNAm values for the two groups. The horizontal gap between the pink and orange lines is the indirect effect of that CpG. Panels A-C illustrate the effect of exposure to maternal psychopathology at 33 months and panel D illustrates the effect of exposure to neighborhood disadvantage at 21 months. **Panel A** shows that average DNAm levels were higher in the exposed group, which correlated higher SMFQ scores. The exposed group in **B** showed higher average DNAm levels, however, SMFQ score decreased as DNAm levels increased. In the exposed group in **C**, average DNAm levels were lower, and higher DNAm levels correlated with higher SMFQ scores. **D** shows how unexposed individuals had higher DNAm levels on average at this CpG, and that higher DNAm correlated with lower SMFQ.

The *positive-negative* pattern was the most common pattern observed, for a total of 23 CpG sites (**Figure 3B**). In this pattern, children with adversity exposure during the sensitive period had increased DNAm, but higher DNAm was correlated with *decreased* depressive symptoms. CpGs exhibiting this pattern explained r=0.01-0.06 of the correlation between childhood adversity and depressive symptoms.

The *negative-positive* pattern was rarer, observed in only 10 CpGs (**Figure 3C**) and indicated that exposed individuals during the sensitive period had reduced DNAm levels at the CpG site, but that higher DNAm levels were correlated with increased SMFQ scores. CpGs exhibiting this pattern explained r=0.01-0.04 of the correlation between childhood adversity and depressive symptoms.

Observed in 18 CpG mediators, the *negative-negative* pattern (**Figure 3D**) indicated that exposed individuals during the sensitive period had reduced DNAm levels at the CpG site, and that higher DNAm levels were correlated with reduced SMFQ scores. CpGs exhibiting this pattern explained r=0.01-0.06 of the correlation between childhood adversity and depressive symptoms.

Detailed information about all 70 sites, including effect estimates, standard errors for effect estimates, strength of each mediator’s indirect effect, whether the CpG is a methylation quantitative trait locus (mQTL), and the nearest gene to that CpG can be found in **Supplemental Table 3**.

#### Biological relevance

To investigate the genetic relevance of our findings, we used the EWAS Toolkit, which is part of the EWAS Atlas developed by Li and colleagues (58). Gene ontology enrichment analysis showed that these 70 sites were linked to genes weakly enriched with 20 biological processes, including chromatin organization and negative epigenetic regulation of gene expression (**Supplemental Figure 3**). However, none of the enrichments survived adjustment for multiple tests (Bonferroni-adjusted p > 0.05). Trait enrichment analysis showed enrichment with CpG sites previously observed to be associated with preterm birth (overlap of 6 CpG sites, Bonferroni-adjusted p < 6e-5), and severe acute malnutrition (overlap of 1 CpG site, Bonferroni-adjusted p < 0.006) (**Supplemental Figure 4**). Analysis of enrichment in regions related to genes, promoters and enhancers, and CpG islands uncovered no major discernable enrichments (**Supplemental Figures 5 and 6**).

### Sensitivity Analyses

We evaluated the usefulness of applying the SLCMA to our data by comparing our results with an approach that only considers an ever-exposed hypothesis. We found that in 6 out of 7 cases, the linear regression model found using SLCMA explained more variability in depressive symptoms than an ever-exposed hypothesis. Furthermore, we ran a mediation analysis using ever-exposed hypotheses instead and found a similar number of mediators, 74 CpG mediators as opposed to the SLCMA’s 70. However, out of these 74, only 6 overlapped with the SLCMA results. Further details and discussion of these results can be found in **Supplemental Materials**.

## Discussion

In this prospective longitudinal study, we identified 70 CpG sites showing evidence of mediating the relationship between exposure to common childhood adversities and depressive symptoms in adolescence. Overall, these CpG sites explained between 10-73% of the correlation observed between adversity and depressive symptoms (the total indirect effect). Interestingly, for most adversities – including sexual/physical abuse, maternal psychopathology, one adult household, and neighborhood disadvantage – we found a negative total indirect (mediating) effect. In other words, for these adversities, changes in DNAm levels at the identified CpG mediators might be *protective* against depressive symptoms. For all remaining adversities, the total indirect effect was positive, suggesting that changes in DNAm levels at the identified CpG mediators worsened depressive symptoms in adolescence. About 1 in 4 of these CpGs were mQTLs, suggesting there are some genetic effects driving these results. These loci were also significantly associated in enrichment analyses with traits connected to childhood adversity, such as preterm birth, a risk factor for negative health outcomes like psychiatric disorders (59) and severe acute malnutrition, a trait closely linked with poverty (60).

We also observed diverse patterns of mediation among these 70 sites, suggesting that if mediation occurs through DNAm, it is unlikely a uniform process. The four patterns – positive-positive, positive-negative, negative-positive, and negative-negative – suggest mediation is likely dependent on numerous factors, including the genomic function of the CpG and its associated gene. This finding supports what we already know: epigenetic adaptation is complex and the effects of childhood adversity on the genome may in fact be plastic and susceptible to intervention (9, 61, 62). However, these findings provide additional evidence that DNAm is not merely a marker of exposure, but rather may be involved in the biological association between childhood adversity and later depressive symptoms.

If replicated in other studies, what might these CpG mediating sites suggest in terms of targets for intervention? These CpG mediating sites help us get closer to understanding the risk pathways that underly the relationship between childhood adversity and depression. In the future, they may help us identify high-risk individuals to target for interventions or prevention strategies. Further investigation into these DNAm mechanisms may help us dissect gene-environment correlations and interactions to assess how much of this risk is preventable and modifiable.

Fundamentally, these results suggest that a negative exposure like childhood adversity can trigger changes in DNAm that then contribute to depressive symptoms. We can therefore deduce that the opposite may also be true – that prevention or intervention methods that reverse these DNAm changes could prevent new onsets of depression or perhaps even reverse or minimize symptoms among people already suffering from depression. Previous studies suggest this reversal may be possible (17, 18, 63). For example, a recent investigation into longitudinal DNAm profiles of soldiers experiencing PTSD found that successful treatment of the disorder with psychotherapy was linked to DNAm changes at 12 differentially methylated regions (DMRs) in various genes (16). Furthermore, the authors found that changes in DNAm at the gene, *ZFP57*, were specifically linked to both the development and remission of PTSD (16). Theirs and others’ findings, coupled with the result patterns we observed, support the urgency to understand these pathways to better inform intervention or prevention strategy.

Our study had several strengths. For one, we used prospective longitudinal data from a unique population-based sample that had repeated measures spanning over a decade. This data structure allowed us to explore how timing and frequency of exposure to childhood adversities played a role in subsequent depressive symptoms. The longitudinal study design also minimized the likelihood that our analysis suffered from reverse causation or recall bias, a limitation often mounted against social epigenetic studies (19). Furthermore, we used a novel mediation analysis technique that allowed for all possible CpG mediators (n=278,586) to have equal consideration for inclusion in the mediation model. Moreover, the CpGs that survived the dimension reduction process were included within the same model, allowing the correlation between CpGs to be factored into calculations. As a result, our estimated mediated effects are more accurate than if the mediators had been considered separately, because the interactions between mediators in this complex biological system are captured (54).

This study was not without limitations, however. As described in **Supplemental Materials**, the Schaid-Sinnwell model involves an iterative process that uses a shrinkage or penalty term to select the optimal number of model parameters as identified by the lowest BIC. A large penalty term selects no mediators while reducing the penalty term increases the number of selected mediators. Because of the relatively small sample size for this study, we opted to balance false-positive and false-negative results by choosing a modest penalty term of 0.2 for all seven adversities. In an ideal scenario, the sample size would be much larger to allow for the possibility to detect greater mediation signal. In addition, our data came from a fairly racially homogenous group (97% of European ancestry). In the future, it would be beneficial to conduct such a study in a more racially diverse group to make these results more generalizable to a broader population.

In summary, our findings provide evidence that changes to DNAm levels might be a consequence of childhood adversity exposure and a potential risk factor for early-adolescent depressive symptoms and provide methylation targets worthy of functional studies. While the mediation analysis technique was exceptional because it allowed us to consider all relevant CpGs in the preliminary steps of our analysis, larger sample sizes and more advanced techniques for analyzing high-dimensional mediators are needed for us to learn more about the biological mechanisms driving depression.

## Data Availability

All data are available by request from the ALSPAC Executive Committee for researchers who meet the criteria for access to confidential data.

http://www.bristol.ac.uk/alspac/researchers/access/

## ACKNOWLEDGEMENTS

We are extremely grateful to all the families who took part in the ALSPAC study, the midwives for their help in recruiting them, and the whole ALSPAC team, which includes interviewers, computer and laboratory technicians, clerical workers, research scientists, volunteers, managers, receptionists and nurses.

