## Supplemental Materials for "DNA methylation partially mediates the relationship between childhood adversity and depressive symptoms in adolescence"

### Details about the reliability of the Short Mood and Feelings Questionnaire (SMFQ) for depressive symptoms

The SMFQ is highly correlated with both questionnaire and interview measures of psychopathology as well as clinician-related diagnoses of depression in children and adolescents (Rhew et al., 2010; Sharp, Goodyer, & Croudace, 2006; Thapar & McGuffin, 1998; Turner, Joinson, Peters, Wiles, & Lewis, 2014). Internal consistency reliability in our sample was very high ($\alpha = 0.78$).

### Details on sociodemographic confounders included as covariates

We controlled for the following sociodemographic confounders measured at child birth: *sex* (0=male, 1=female); *child race/ethnicity* (0=non-White, 1=White); *birthweight*; *number of previous pregnancies (parity)* (0-3+); *maternal age*; *highest level of maternal education* (1=less than O-level, 2=O-level, 3=A-level, 4=degree or above); *sustained maternal smoking during pregnancy* (0=non-smoker, 1=smoker in two or more trimesters, including the third trimester).

### Childhood adversity exposure variable parameterization in the structured life course modeling approach (SLCMA)

The SLCMA analysis we performed required three categories of exposure variables. To test for sensitive periods, a binary variable with a value of 1 for exposed or 0 for unexposed was assigned at each measurement timepoint. To test the accumulation hypothesis, a variable was created by summing the binary measurement points for each adversity. Finally, to test the recency hypothesis, each binary measurement point was multiplied by the child's age in years at that timepoint. This gave greater weight to more recent exposures to adversity. These weighted values then were summed to create the recency hypothesis variable.

**CpG reduction techniques**

We used pruning and sure independence screening (SIS) to reduce the number of DNAm mediators to a viable number, $q = \lfloor (n - 3 - 1)/2 \rfloor$, where n equals the analytic sample size for each adversity. This value of $q$ was chosen because it maximized the number of potential mediation sites we could consider in each model, while remaining below the number of parameters we could estimate for each sample size.

We started by subtracting three from $n$ to accommodate the estimation of the direct effect of exposure on outcome, $\partial$ as well as the variances of the exposure and outcome. An additional one was subtracted to ensure that the final number of parameters estimated was at least one less than the sample size. Finally, we divided by two to accommodate that each potential mediator required the estimation of both an $\alpha$ and a $\beta$ parameter.

<u>Pruning</u>

Given that our analyses were not stratified by sex, we removed CpGs located in sex chromosomes from consideration. We also removed non-variable CpGs (i.e., CpGs whose methylation levels varied by less than 5% across the sample), as these CpGs may not provide much insight into the underlying mechanisms of the childhood adversity-adolescent depression relationship. This left us with 278,586 probes for downstream analyses.

Sure independence screening (SIS)

We then made use of a method proposed by Fan & Lv to reduce our probes to the pre-specified $q$, selecting the top sites by ranking the remaining 278,586 probes by the absolute value of their marginal correlation with each adversity and SMFQ score (Fan & Lv, 2008; Schaid & Sinnwell, 2020). This calculation took the form:

$$\left|cor\left(x, m_j\right) \cdot cor\left(m_j, y\right)\right|, \quad (1)$$

where x is the adversity exposure, $m_j$ is a given CpG site where $j = 1, \ldots, 278{,}586$, and $y$ is SMFQ score. Thus, the assumption is: CpGs that show stronger correlations with both the exposure and the outcome are more likely to be mediators than CpGs with lower or no correlation with exposure and outcome. Therefore, we focused on the CpGs showing the greatest likelihood of being mediators and selected the top $q$ sites to include in our model.

Shrinkage parameter

Once the top $q$ mediation sites were entered into the SEM, the SEM's iterative algorithm identified the CpG sites showing the strongest mediation signal by systematically working through a grid of values for the shrinkage parameter, $\lambda$, beginning with 1 (the most stringent penalty) and gradually reducing to zero (no penalty) by 0.1 increments. The iterative selection of a penalty parameter is necessary to identify the number of model parameters that result in the lowest Bayesian Information Criterion (BIC).

**Sensitivity Analyses**

We ran sensitivity analyses to test the effects of using the SLCMA over a simple, binary ever-exposed hypothesis. In the first, we compared the $R^2$ values for a linear regression model containing confounding covariates and the SLCMA-selected exposure variable to a linear regression model containing confounding covariates and the ever-exposed variable. In all but one case, where differences between the two values were negligible, the SLCMA explained more variability in the outcome. This suggests that these models do a better job of explaining the relationship between adversity and depressive symptoms than simplistic ever-exposed models.

We also tested the ever-exposed hypothesis The ever-exposed analysis yielded a similar number of mediators – 74 versus the SLCMA's 70 – however, only 6 of these 74 CpGs overlapped with the 70 CpGs found using the SLCMA. The lack of overlap suggests that, in this instance, these forms of exposure (ever-exposed versus sensitive period) are not interchangeable at a biological level. If that is the case, it would have major implications for DNAm mediation analysis, warranting more thoughtful and precise definitions of exposures before conducting the analysis.

**Supplemental Table 1**. **Description of seven childhood adversity measures included as exposures in the current study.**

| Adversity | Respondent | Instrument or questionnaire items | Exposure definition | Time points of assessment |
|---|---|---|---|---|
| **Caregiver physical or emotional abuse** | Mother and partner | 1) your partner was physically cruel to your children; 2) you were physically cruel to your children; 3) your partner was emotionally cruel to your children; 4) you were emotionally cruel to your children | Children were coded as having been exposed if the mother, partner, or both responded affirmatively to at least one item. | 8 months, 1.75 years, 2.75 years, 4 years, 5 years, and 6 years |
| **Sexual or physical abuse** | Mother | An item asking whether the child had been exposed to either sexual or physical abuse from anyone | Children were coded as exposed if the mother responded affirmatively. | 1.5 years, 2.5 years, 3.5 years, 4.75 years, 5.75 years, and 6.75 years |
| **Maternal psychopathology** | Mother | 1) the Crown-Crisp Experiential Index (CCEI), assessing anxiety and depression; 2) the Edinburgh Postnatal Depression Scale (EPDS); and 3) a question asking about suicide attempts in the past 1.5 years | Consistent with cut-offs used in prior ALSPAC studies and elsewhere, children were coded as exposed if one or more of the following criteria was met: 1) CCEI depression score > 9 and/or anxiety score > 10; 2) EPDS score > 12; or 3) a suicide attempt reported | 8 months, 1.75 years, 2.75 years, 5 years, and 6 years of age |
| **One adult in the household** | Mother | An item asking about the number of adults over the age of 18 years living in the household | Children were coded as exposed if only one adult was lived in the household. | 8 months, 1.75 years, 2.75 years, 4 years, and 7 years |

| | | | | |
|---|---|---|---|---|
| **Family instability** | Mother | Child had 1) been taken into care; 2) been separated from their mother for two or more weeks; 3) been separated from their father for two or more weeks; or 4) acquired a new parent. | Children were coded as exposed if mothers responded affirmatively to at least two of these events at a single time point. | 1.5 years, 2.5 years, 3.5 years, 4.75 years, 5.75 years, and 6.75 years |
| **Financial hardship** | Mother | The family had difficulty affording the following: 1) items for the child; 2) rent or mortgage; 3) heating; 4) clothing; 5) food. Each of the 5 items was coded on a Likert-type scale (1=not difficult; 2=slightly difficult; 3=fairly difficult; 4=very difficult) | Children were coded as exposed if mothers reported at least "fairly difficult" (corresponding to 3 or higher) for three or more items at a single time point. | 8 months, 1.75 years, 2.75 years, 5 years, and 7 years |
| **Neighborhood disadvantage** | Mother | There were problems in the neighborhood: 1) noise from other homes; 2) noise from the street; 3) garbage on the street; 4) dog dirt; 5) vandalism; 6) worry about burglary; 7) mugging; and 8) disturbance from youth. Response options to each item were: 2=serious problem, 1=minor problem, 0=not a problem or no opinion. | A sum score ranging from 0-16 was created based on responses to each item. Children were coded as exposed if they had scores greater than or equal to 8. | 1.75 years, 2.75 years, 5 years, and 7 years |

**Supplemental Table 2. Descriptive statistics of the analytic sample.**

| Variable | Level | Analytic Sample (AS) N = 675 | ARIES N = 970 | *p*-value (AS to ARIES) | ALSPAC N = 15,646 | *p*-value (ARIES to ALSPAC) |
|---|---|---|---|---|---|---|
| Sex (%) | Female | 325 (48.1) | 490 (50.5) | 0.371 | 7152 (48.7) | 0.281 |
| | Male | 350 (51.9) | 480 (49.5) | | 7542 (51.3) | |
| Race (%) | Non-white | 22 (3.3) | 27 (2.9) | 0.784 | 611 (5.1) | 0.004 |
| | White | 653 (96.7) | 906 (97.1) | | 11488 (94.9) | |
| Birthweight (g) (%) | < 3000 | 94 (13.9) | 128 (13.5) | 0.943 | 2760 (20.0) | <0.001 |
| | 3000 - 3499 | 243 (36.0) | 342 (36.0) | | 4924 (35.7) | |
| | 3500 - 3999 | 229 (33.9) | 334 (35.2) | | 4382 (31.8) | |
| | 4000 | 109 (16.1) | 146 (15.4) | | 1735 (12.6) | |
| Mother's birth age (mean (SD)) | | 30.01 (4.20) | 29.57 (4.46) | 0.045 | 28.00 (4.96) | <0.001 |
| Previous number of pregnancies (%) | 0 | 308 (45.6) | 435 (46.5) | 0.96 | 5800 (44.7) | 0.015 |
| | 1 | 259 (38.4) | 348 (37.2) | | 4550 (35.0) | |
| | 2 | 84 (12.4) | 117 (12.5) | | 1860 (14.3) | |
| | 3+ | 24 (3.6) | 36 (3.8) | | 772 (5.9) | |
| Education (%) | CSE/Vocational /None | 82 (12.1) | 148 (15.6) | 0.235 | 3735 (30.0) | <0.001 |
| | O Level | 230 (34.1) | 324 (34.1) | | 4303 (34.6) | |
| | A level | 213 (31.6) | 281 (29.6) | | 2795 (22.5) | |
| | University Degree | 150 (22.2) | 196 (20.7) | | 1603 (12.9) | |
| Smoked during pregnancy (%) | No | 612 (90.7) | 818 (89.2) | 0.384 | 9565 (78.8) | <0.001 |
| | Yes | 63 (9.3) | 99 (10.8) | | 2577 (21.2) | |
| Short Mood and Feelings Questionnaire (SMFQ), mean age 10.6 (mean (SD)) | | 3.70 (3.27) | 3.83 (3.32) | 0.431 | 4.04 (3.51) | 0.086 |

Caregiver physical/emotional abuse analytic sample used as an example. Analytic sample did not statistically ($\alpha = 0.05$) differ from total ARIES sample except for a slight discrepancy in SMFQ scores. P-values are from the chi-squared test (categorical variables) or a two-sample t-test (continuous variables).

**Supplemental Table 3. Results from mediation analysis examining DNAm at age 7 as a mediator of the relationship between exposure to childhood adversity from ages 0-7 and depressive symptoms at age 10.6.**

| Adversity | Theoretical Model | CpG | $\alpha$[*] | SE[†] ($\alpha$) | $\beta$[#] | SE ($\beta$) | Indirect Effect | CI[1] | *P*-value | Nearest gene |
|---|---|---|---|---|---|---|---|---|---|---|
| Caregiver Physical/ Emotional Abuse | Very Early Childhood | | | | | | | | | |
| | | **cg23751110[2]** | **-0.101** | **0.038** | **-0.152** | **0.038** | **0.015** | **(0.003, 0.031)** | **0.006** | **SLIT2** |
| | | **cg06804625[2]** | **-0.102** | **0.038** | **-0.115** | **0.038** | **0.012** | **(0.002, 0.025)** | **0.008** | **PSRC1** |
| | | **cg21089584[2]** | **0.099** | **0.037** | **-0.117** | **0.037** | **-0.012** | **(-0.025, -0.002)** | **0.009** | **JARID2** |
| | | **cg12343929** | **0.096** | **0.039** | **0.121** | **0.039** | **0.012** | **(0.002, 0.025)** | **0.013** | **WDR90** |
| | | **cg03965496** | **0.084** | **0.037** | **0.115** | **0.037** | **0.01** | **(0.001, 0.022)** | **0.023** | **TRNA_Asn** |
| Sexual/Physical Abuse (by anyone) | Very Early Childhood | | | | | | | | | |
| | | **cg24622544[2]** | **-0.113** | **0.039** | **-0.141** | **0.037** | **0.016** | **(0.004, 0.032)** | **0.003** | **POP5** |
| | | **cg10310274** | **-0.112** | **0.039** | **-0.116** | **0.037** | **0.013** | **(0.003, 0.028)** | **0.005** | **UCK1** |
| | | **cg26518628[2]** | **0.105** | **0.039** | **-0.111** | **0.037** | **-0.012** | **(-0.025, -0.002)** | **0.008** | **7SK** |
| | | **cg00958217** | **-0.097** | **0.039** | **-0.127** | **0.037** | **0.012** | **(0.002, 0.027)** | **0.013** | **THAP3** |
| | | **cg26786980** | **-0.087** | **0.039** | **-0.101** | **0.037** | **0.009** | **(0.001, 0.021)** | **0.028** | **RARB** |
| | | **cg01973483** | **0.077** | **0.039** | **-0.145** | **0.037** | **-0.011** | **(-0.026, 0)** | **0.045** | **PITPNM2** |
| Maternal Psychopathology | Very Early Childhood | | | | | | | | | |
| | | **cg24059871[3]** | **0.096** | **0.039** | **0.122** | **0.037** | **0.012** | **(0.002, 0.025)** | **0.016** | **POP4** |
| | | **cg16292933[2]** | **-0.089** | **0.039** | **0.097** | **0.037** | **-0.009** | **(-0.02, -0.001)** | **0.03** | **SLC4A8** |
| | | **cg19642007** | **0.087** | **0.039** | **0.109** | **0.037** | **0.009** | **(0.001, 0.022)** | **0.031** | **TNNT3** |
| | | cg10953317[3] | 0.064 | 0.039 | -0.158 | 0.037 | -0.01 | (-0.025, 0.002) | 0.105 | CD300A |
| | | cg00300275 | 0.062 | 0.039 | -0.138 | 0.037 | -0.009 | (-0.022, 0.002) | 0.111 | U80764 |
| | | cg06451157 | 0.067 | 0.039 | -0.081 | 0.039 | -0.005 | (-0.016, 0.001) | 0.118 | HLA-H |
| | | cg07308232 | 0.086 | 0.039 | -0.059 | 0.038 | -0.005 | (-0.015, 0.001) | 0.148 | C7orf50 |
| | | cg21665774 | 0.071 | 0.039 | -0.069 | 0.05 | -0.005 | (-0.016, 0.002) | 0.225 | KIAA0355 |
| | | cg18571112 | -0.083 | 0.039 | 0.058 | 0.05 | -0.005 | (-0.017, 0.003) | 0.275 | SFMBT1 |
| One Adult in Household | Early Childhood | | | | | | | | | |
| | | **cg22239534[3]** | **-0.093** | **0.039** | **0.174** | **0.037** | **-0.016** | **(-0.032, -0.003)** | **0.015** | **AK123632** |
| | | **cg06456365** | **0.086** | **0.039** | **-0.131** | **0.036** | **-0.011** | **(-0.025, -0.001)** | **0.026** | **RFPL4B** |
| | | **cg21079003[2]** | **-0.084** | **0.039** | **0.119** | **0.036** | **-0.01** | **(-0.023, -0.001)** | **0.03** | **RGMA** |
| | | **cg20930329** | **0.078** | **0.039** | **0.122** | **0.036** | **0.009** | **(0, 0.022)** | **0.043** | **AY748447** |
| | | cg03269218[2] | -0.076 | 0.039 | 0.114 | 0.036 | -0.009 | (-0.021, 0) | 0.051 | BC043227 |
| | | cg26078436 | -0.08 | 0.039 | -0.1 | 0.041 | 0.008 | (0, 0.02) | 0.054 | HBBP1 |
| | | cg22255773 | 0.082 | 0.039 | -0.081 | 0.037 | -0.007 | (-0.017, 0) | 0.06 | LOC100132707 |

| | | | | | | | | | | |
|---|---|---|---|---|---|---|---|---|---|---|
| | | cg00901198 | 0.078 | 0.039 | -0.084 | 0.037 | -0.007 | (-0.017, 0) | 0.064 | BAI2 |
| | | cg09191574 | 0.048 | 0.039 | -0.098 | 0.04 | -0.005 | (-0.015, 0.003) | 0.229 | DIO2 |
| | | cg03800296 | -0.075 | 0.039 | -0.027 | 0.041 | 0.002 | (-0.005, 0.01) | 0.542 | TRNA_Gln |
| Family Instability | Early Childhood | | | | | | | | | |
| | | **cg27200630** | **0.124** | **0.039** | **-0.171** | **0.038** | **-0.021** | **(-0.039, -0.007)** | **0.002** | **PIK3CB** |
| | | **cg21011883** | **0.125** | **0.039** | **0.116** | **0.037** | **0.014** | **(0.004, 0.029)** | **0.003** | **L1TD1** |
| | | **cg26299079**[2] | **0.114** | **0.039** | **-0.124** | **0.037** | **-0.014** | **(-0.029, -0.003)** | **0.003** | **BTBD16** |
| | | **cg26389281** | **-0.107** | **0.039** | **-0.148** | **0.037** | **0.016** | **(0.004, 0.032)** | **0.006** | **ABR** |
| | | **cg21305041** | **-0.105** | **0.039** | **-0.136** | **0.048** | **0.014** | **(0.002, 0.031)** | **0.011** | **SH3BGRL2** |
| | | **cg16087263** | **0.119** | **0.039** | **0.091** | **0.037** | **0.011** | **(0.001, 0.024)** | **0.014** | **PLA2G2F** |
| | | cg25513610 | 0.156 | 0.039 | 0.08 | 0.043 | 0.013 | (0, 0.029) | 0.061 | CD83 |
| | | cg22839587 | -0.113 | 0.039 | -0.054 | 0.051 | 0.006 | (-0.005, 0.02) | 0.29 | DPYSL3 |
| Financial Stress | Early Childhood | | | | | | | | | |
| | | **cg10953317**[2,3] | **-0.159** | **0.039** | **-0.115** | **0.036** | **0.018** | **(0.006, 0.035)** | **0.001** | **CD300A** |
| | | **cg02674870**[2] | **-0.123** | **0.04** | **-0.136** | **0.036** | **0.017** | **(0.005, 0.033)** | **0.0019** | **Mir_598** |
| | | **cg21202551**[2] | **-0.105** | **0.04** | **0.109** | **0.035** | **-0.011** | **(-0.025, -0.002)** | **0.009** | **MIR4710** |
| | | **cg20777315** | **0.101** | **0.04** | **0.124** | **0.036** | **0.012** | **(0.002, 0.026)** | **0.0134** | **LOC100505536** |
| | | **cg23462687** | **0.088** | **0.04** | **-0.15** | **0.035** | **-0.013** | **(-0.028, -0.001)** | **0.0281** | **HDAC4** |
| | | **cg00188315** | **-0.085** | **0.04** | **-0.124** | **0.036** | **0.011** | **(0.001, 0.024)** | **0.0335** | **LOC285501** |
| | | **cg11293312** | **-0.082** | **0.04** | **0.101** | **0.036** | **-0.008** | **(-0.02, 0)** | **0.0437** | **IZUMO1** |
| | | cg22239534[2,3] | -0.077 | 0.04 | 0.114 | 0.036 | -0.009 | (-0.021, 0) | 0.0567 | AK123632 |
| | | cg11738723 | 0.078 | 0.04 | -0.101 | 0.04 | -0.008 | (-0.02, 0) | 0.0611 | AX747193 |
| | | cg07118000[2] | -0.078 | 0.04 | -0.088 | 0.035 | 0.007 | (0, 0.018) | 0.0624 | GPR124 |
| | | cg04347379[2] | 0.07 | 0.04 | 0.144 | 0.037 | 0.01 | (-0.001, 0.024) | 0.077 | HEATR2 |
| | | cg16515600 | 0.123 | 0.04 | 0.064 | 0.037 | 0.008 | (-0.001, 0.02) | 0.0843 | PHTF1 |
| | | cg02389555 | -0.068 | 0.04 | 0.128 | 0.036 | -0.009 | (-0.022, 0.001) | 0.0872 | TRIM27 |
| | | cg06820822[2] | 0.07 | 0.04 | -0.083 | 0.037 | -0.006 | (-0.016, 0.001) | 0.1033 | C7orf62 |
| | | cg03033975 | 0.057 | 0.04 | 0.134 | 0.037 | 0.008 | (-0.003, 0.021) | 0.1568 | GTPBP5 |
| | | cg16701559 | -0.075 | 0.04 | -0.048 | 0.039 | 0.004 | (-0.002, 0.013) | 0.2726 | RPP21 |
| | | cg23798471 | 0.071 | 0.04 | -0.027 | 0.041 | -0.002 | (-0.01, 0.004) | 0.5347 | SNORA27 |
| Neighborhood Disadvantage | Very Early Childhood | | | | | | | | | |
| | | **cg18604823** | **0.12** | **0.039** | **-0.192** | **0.036** | **-0.023** | **(-0.042, -0.008)** | **0.002** | **GALP** |
| | | **cg13003513** | **-0.095** | **0.039** | **0.107** | **0.036** | **-0.01** | **(-0.023, -0.001)** | **0.016** | **CARD11** |
| | | **cg11611320** | **0.104** | **0.039** | **-0.099** | **0.04** | **-0.01** | **(-0.024, -0.001)** | **0.021** | **LOC401242** |
| | | **cg15027300** | **0.086** | **0.039** | **-0.115** | **0.035** | **-0.01** | **(-0.022, -0.001)** | **0.029** | **GGN** |
| | | **cg08470892**[2] | **0.088** | **0.039** | **0.105** | **0.036** | **0.009** | **(0.001, 0.021)** | **0.029** | **TPSD1** |
| | | **cg20262683** | **0.085** | **0.039** | **0.09** | **0.035** | **0.008** | **(0, 0.019)** | **0.039** | **NPTX2** |
| | | cg14223671[2] | 0.074 | 0.039 | -0.081 | 0.035 | -0.006 | (-0.016, 0) | 0.075 | PRR25 |

| | | | | | | | | |
|---|---|---|---|---|---|---|---|---|
| cg24059871[3] | 0.069 | 0.039 | 0.121 | 0.035 | 0.008 | (-0.001, 0.02) | 0.078 | POP4 |
| cg27423959 | 0.066 | 0.039 | -0.134 | 0.035 | -0.009 | (-0.022, 0.001) | 0.087 | C3orf56 |
| cg24738171 | 0.059 | 0.039 | 0.129 | 0.038 | 0.008 | (-0.002, 0.02) | 0.127 | HGS |
| cg05931366 | -0.057 | 0.039 | -0.137 | 0.037 | 0.008 | (-0.003, 0.021) | 0.149 | LINC00266-1 |
| cg08073133 | 0.122 | 0.039 | 0.051 | 0.037 | 0.006 | (-0.002, 0.018) | 0.167 | SERPINE2 |
| cg23119063[2] | 0.099 | 0.039 | 0.041 | 0.037 | 0.004 | (-0.003, 0.014) | 0.272 | Mir_320 |
| cg26595256 | 0.083 | 0.039 | 0.04 | 0.037 | 0.003 | (-0.002, 0.012) | 0.294 | TRIO |
| cg01439119 | -0.075 | 0.039 | -0.039 | 0.039 | 0.003 | (-0.003, 0.011) | 0.36 | POLR3B |

[*] $\alpha$= effect of adversity on DNAm at given CpG; [#] $\beta$= effect of DNAm at CpG on depressive symptoms; [2] CpG site is a methylation quantitative trait locus (mQTL), meaning a locus with DNA methylation levels that are influenced by genetics (19 total mGTLs present); [3] Duplicate CpG sites (cg10953317, cg22239534, cg24059871) appearing in two different adversities each; [†] SE = standard error; [1] CI = confidence interval; **bolded** rows indicate statistically significant results at $p<0.05$ level.

**Supplemental Figure 1. Prevalence of exposures within each adversity's analytic sample**.

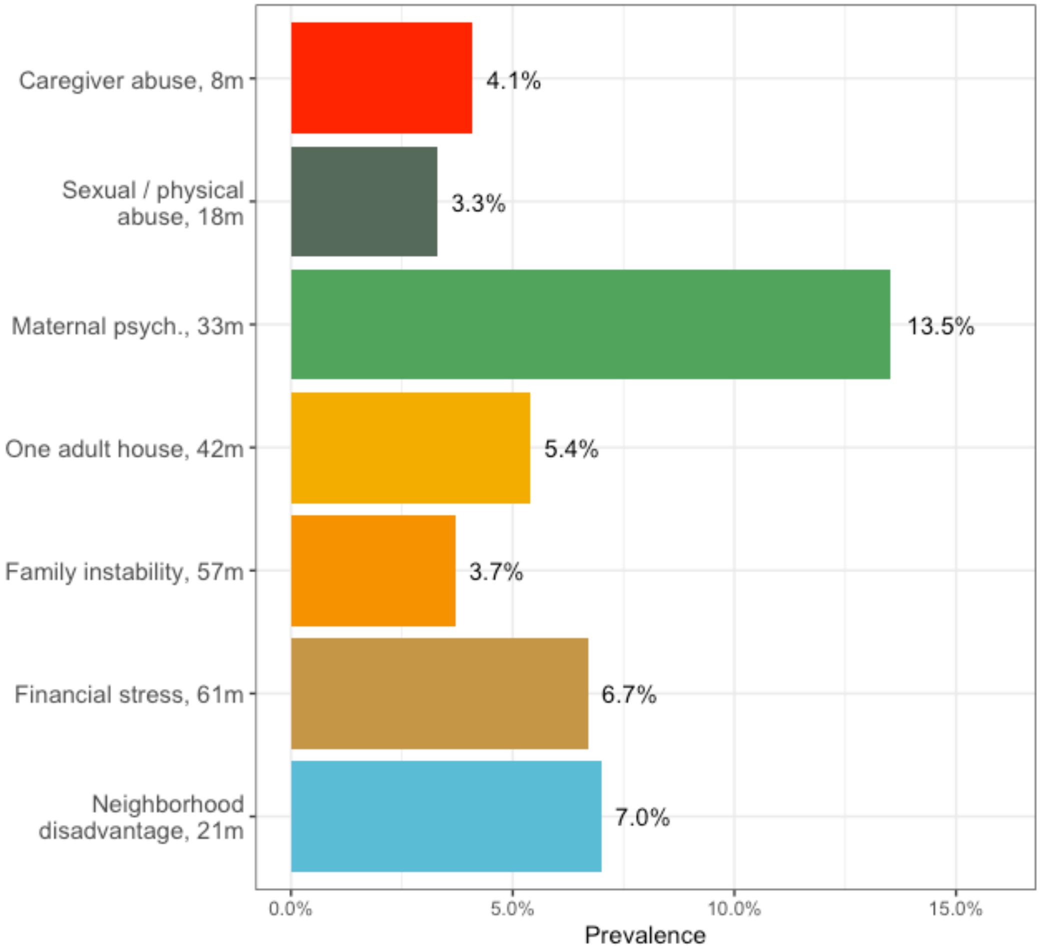


Prevalence of adversity exposure within each analytic sample ranged from 3.3% in sexual/physical abuse (by anyone) to 13.5% in maternal psychopathology.

**Supplemental Figure 2. Single and multiple mediator structures**.

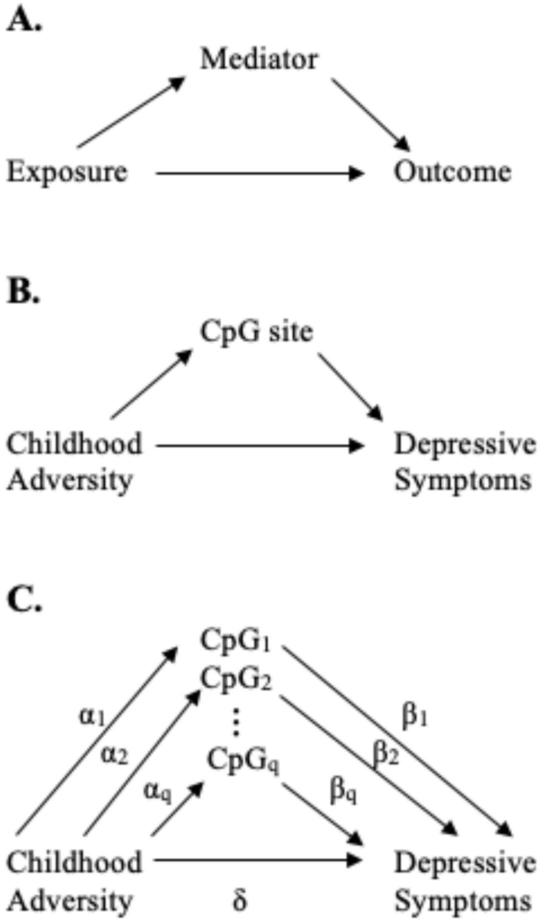


Directed acyclic graph (DAG) **1A.** below shows a general exposure-mediator-outcome relationship with a single mediator and then in **1B.** within the context of our study. **1C.** shows a simplified version of our multiple mediator analysis where q represents the total number of mediators considered in the analysis after sure independence screening. Key: CpG = DNA region where a cytosine nucleotide is followed by a guanine nucleotide; $\alpha_i$ = effect estimate of childhood adversity on $CpG_i$; $\beta_i$ = effect estimate of $CpG_i$ DNA methylation on depressive symptoms

**Supplemental Figure 3. Gene ontology enrichment of mediating loci (n=70)**.

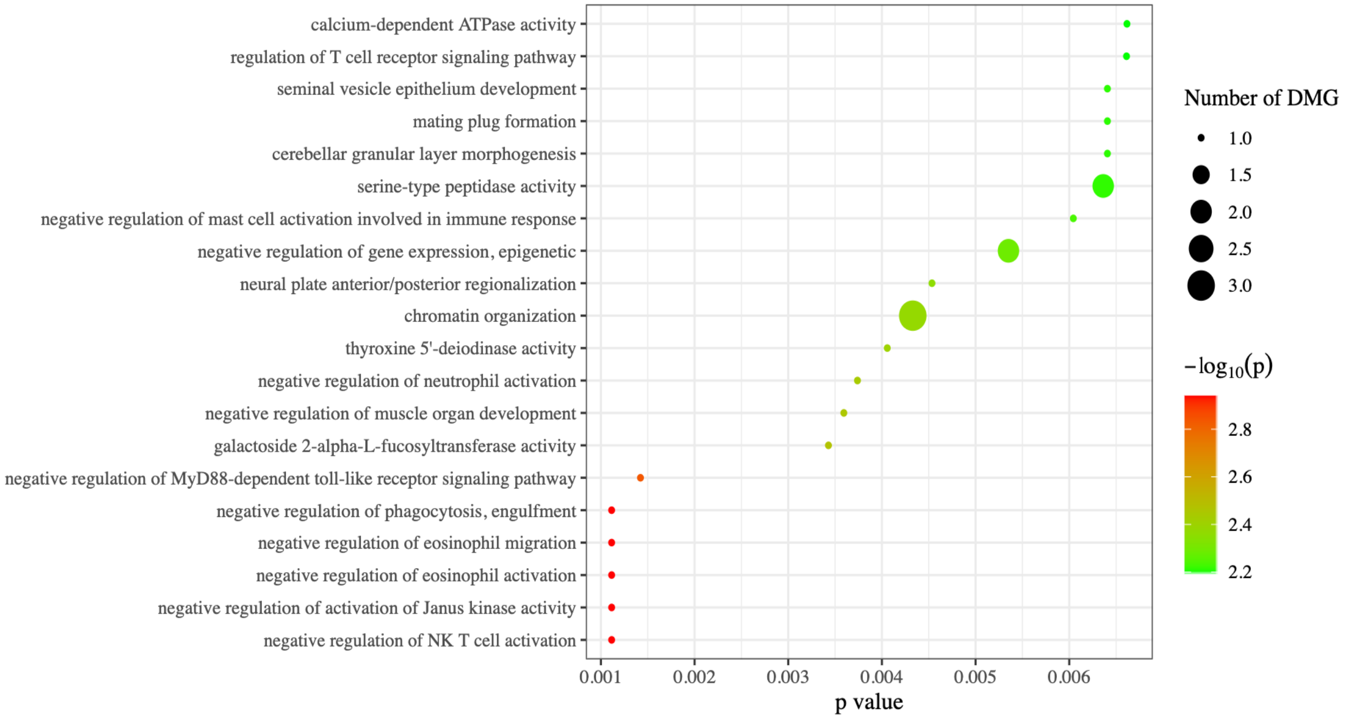


Source: EWAS Atlas. Results of a gene ontology enrichment analysis show 20 biological processes affiliated with these 70 mediating loci. The size of the points reflects the number of differentially methylated genes (DMGs) associated with each process and the color indicates the statistical significance of this association. Figure shows that these 70 sites were linked to genes weakly enriched with 20 biological processes. None of the enrichments survived adjustment for multiple tests (Bonferroni-adjusted $p > 0.05$).

**Supplemental Figure 4. Trait enrichment of mediating loci (n=70).**

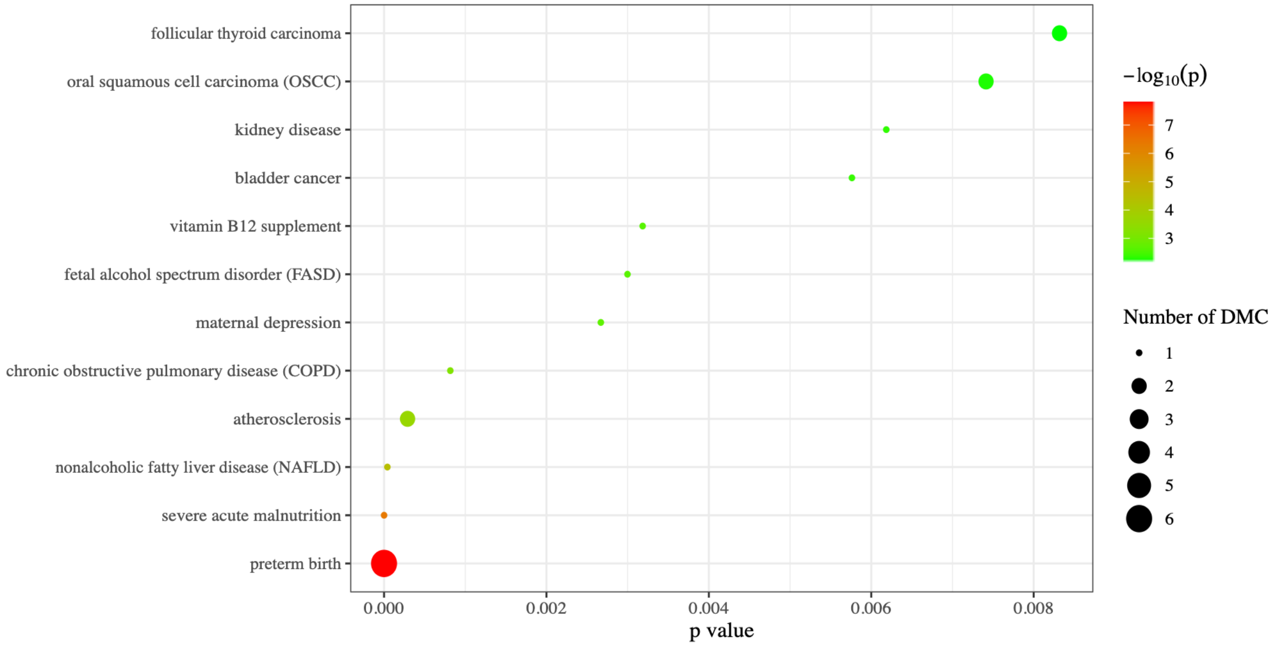


Source: EWAS Atlas. Results of a trait enrichment analysis show 12 traits affiliated with these 70 mediating loci. The size of the points reflects the number of differentially methylated CpGs (DMCs) associated with each process and the color indicates the statistical significance of this association. Trait enrichment analysis showed that enrichment with CpG sites previously observed to be associated with preterm birth (overlap of 6 CpG sites, Bonferroni-adjusted $p < 6e-5$), and severe acute malnutrition (overlap of 1 CpG site, Bonferroni-adjusted $p < 0.006$).

**Supplemental Figure 5. Genomic features for all mediators and mediator patterns**.

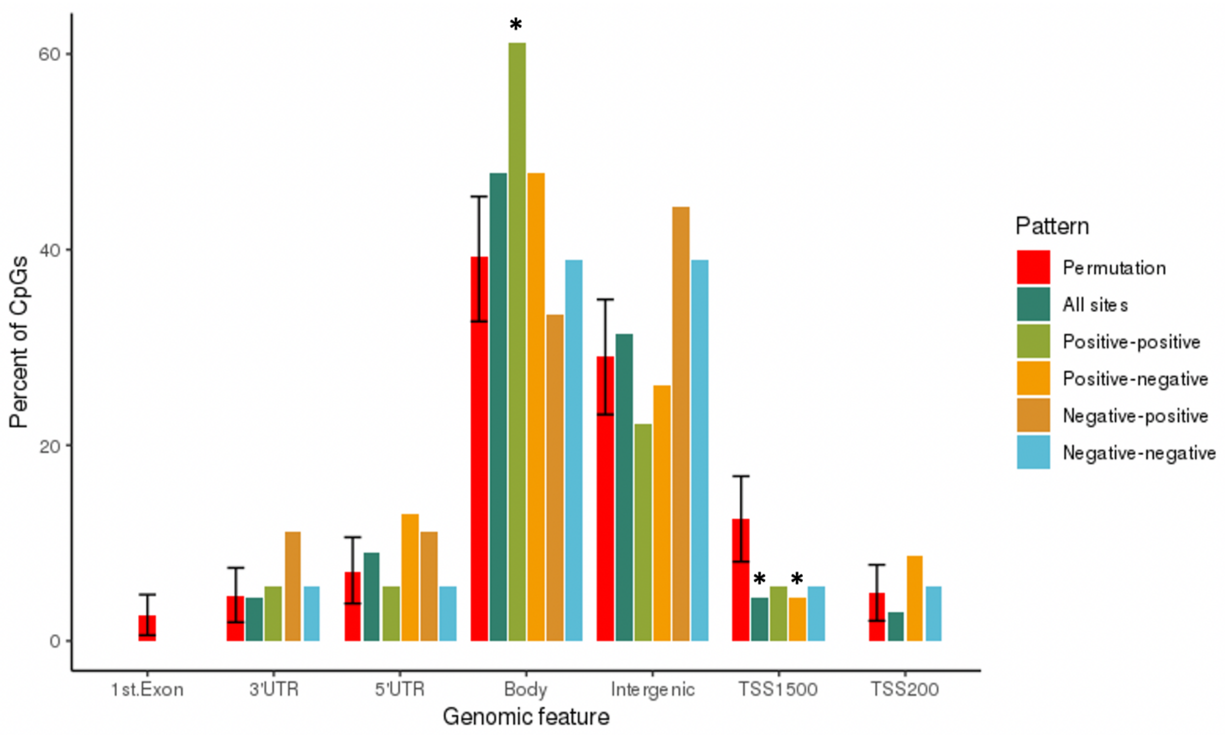


The graph above depicts where the selected mediators fall in the genome. The x-axis represents the seven categories of genomic features and the y-axis measures the percent of CpGs that fall within each category. Red bars represent permuted values, which are the percent of CpGs for each genomic feature that we would expect by chance. Dark green bars represent the percent of CpGs that fall into each pattern for all 67 unique mediators. The remaining bars separate the CpG sites by mediator pattern. A * indicates the amount was greater than random chance at $p<0.05$.

**Supplemental Figure 6. Genomic locations of mediating loci (n=70) compared to all sites tested (n=278,586).**

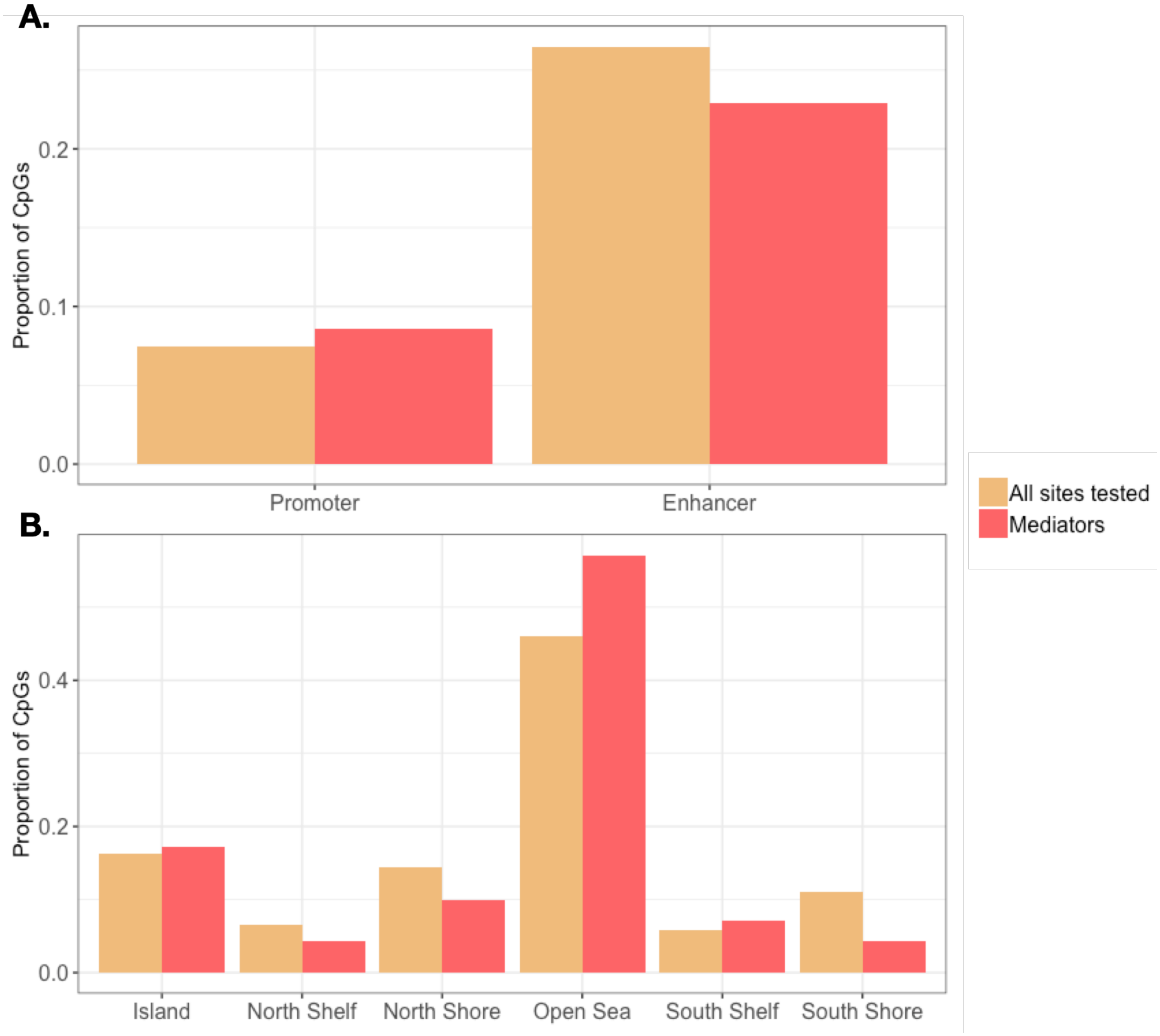


**A.** Compared to all tested sites, mediating loci showed less enrichment in enhancer regions ($\chi^2$=0.307; p=0.58) and more enrichment in promoter regions ($\chi^2$=0.014; p=0.91). **B.** Mediating loci differed in terms of their location relation to CpG islands, showing higher enrichment in Open Sea regions and CpG islands, while showing decreased enrichment in southern shore regions compared to all sites ($\chi^2$= 6.503; p= 0.26).
